# Body composition as a biomarker for assessing future lung cancer risk

**DOI:** 10.1101/2024.10.14.24315477

**Authors:** Jing Wang, Joseph K. Leader, Xin Meng, Tong Yu, Renwei Wang, James Herman, Jian-min Yuan, David Wilson, Jiantao Pu

**Affiliations:** Department of Radiology, School of Medicine, University of Pittsburgh, Pittsburgh, PA 15213, USA; Department of Bioengineering, University of Pittsburgh, Pittsburgh, PA 15213, USA; Cancer Epidemiology and Prevention Program, UPMC Hillman Cancer Center, Pittsburgh, PA 15232, USA; Division of Oncology, Department of Medicine, University of Pittsburgh, PA 15213, USA; Department of Epidemiology, School of Public Health, University of Pittsburgh, Pittsburgh, PA 15213, USA; Division of Pulmonary, Allergy and Critical Care Medicine, Department of Medicine, School of Medicine, University of Pittsburgh, Pittsburgh, PA 15213, USA; Department of Ophthalmology, School of Medicine, University of Pittsburgh, PA 15213, USA

**Author notes:** Corresponding authors and guarantors of the entire manuscript: Jiantao Pu, PhD, Contact E-mail address, Mailing address: 3240 Craft Place, Pittsburgh, PA 15213. Disclosure Statement: The authors have no conflicts of interest to declare. This work has been submitted to Radiology: Imaging Cancer for possible publication. Copyright may be transferred without notice, after which time this version may no longer be accessible.

**Keywords:** Biomarkers, Body composition, Lung cancer, Time-to-event analysis

## Abstract

**Purpose:** To investigate if body composition is a biomarker for assessing the risk of developing lung cancer.

**Materials and Methods:** Low-dose computed tomography (LDCT) scans from the Pittsburgh Lung Screening Study (PLuSS) (n=3,635, 22 follow-up years) and NLST-ACRIN (n=16,435, 8 follow-up years) cohorts were used in the study. Artificial intelligence (AI) algorithms were developed to automatically segment and quantify subcutaneous adipose tissue (SAT), visceral adipose tissue (VAT), intramuscular adipose tissue (IMAT), skeletal muscle (SM), and bone. Cause-specific Cox proportional hazards models were used to evaluate the hazard ratios (HRs). Standard time-dependent receiver operating characteristic (ROC) analysis was used to evaluate the prognostic ability of different models over time.

**Results:** The final composite models were formed by seven variables: age (HR=1.20), current smoking status (HR=1.59), bone volume (HR=1.79), SM density (HR=0.29), IMAT ratio (HR=0.33), IMAT density (HR=0.56), and SAT volume (HR=0.56). The models trained on the PLuSS cohort achieved a mean AUC of 0.76 (95% CI: 0.74-0.79) over 21 follow-up years and 0.70 (95% CI: 0.66-0.74) over the first 7 follow-up years for predicting lung cancer development within the PLuSS cohort. In contrast, models trained on the PLuSS cohort alone, as well as in combination with the NLST cohorts, achieved an AUC ranging from 0.61 to 0.68 in the NLST cohort over a 7-year follow-up period.

**Conclusion:** Body composition assessed on LDCT is a significant predictor of lung cancer risk and could improve the effectiveness of LDCT lung cancer screening by optimizing the screening eligibility and frequency.

**Summary statement:** Body composition assessed on LDCT is a significant predictor of lung cancer risk and could improve the effectiveness of LDCT lung cancer screening by optimizing the screening eligibility and frequency.

**Key Points:**

1. This study unveils the significant associations between body tissues and lung cancer risk.
2. The prediction models based on body composition alone, as well as the combination of demographics and body composition features can effectively identify patients at higher risk of developing lung cancer.

## INTRODUCTION

Lung cancer is the leading cause of cancer-related deaths worldwide^1^. Several large studies, such as the National Lung Screening Trial (NLST)^2–4^ and the NELSON study^5^, have confirmed that screening with low-dose computed tomography (LDCT) scans can significantly reduce lung cancer-related deaths compared to chest radiography. These outcomes ultimately led to the approval and reimbursement for lung cancer screening using LDCT scans. The current eligibility criteria for lung cancer screening with LDCT are primarily based on age and smoking history, specifically, adults aged 50–80 years with a 20-pack-year or more smoking history who either smoked or quit in the last 15 years^6^. Among the screening eligible populations, only a small percentage (<4%) of screened individuals have or will develop lung cancer^2, 3^. In the NLST, prevalent and first-incident lung cancers were detected in only 1.1 % and 0.7% of the patients screened, respectively, with positive predictive values being 3.8% and 2.4%, respectively ^2, 3^. Among the 53,454 patients who smoke or previously smoked enrolled in NLST, only 3.85% (2,058 subjects) were diagnosed with lung cancer in the study period (2002–2010). Therefore, in the current lung cancer screening paradigm, a significant majority of the screened individuals will not develop lung cancer but may be unnecessarily exposed to potentially harmful radiation or other procedures (e.g., biopsy). Moreover, current lung cancer screening relies on the detection and evaluation of lung nodules ^7–9^. However, lung nodules are only detected in a modest percentage of screened individuals, and 96% of the screen-detected nodules are false positives (non-cancerous)^2^. Therefore, it is an unmet clinical need for improving lung cancer screening by identifying individuals at high risk of developing lung cancer using an approach that goes beyond the current eligibility criteria and does not solely rely on the presence of lung nodules. For those at low risk of developing lung cancer, less frequent LDCT-based screening (e.g., every 2-3 years rather than yearly) may be more appropriate.

Available methods for assessing lung cancer risk primarily rely on clinical and demographic variables ^10^ ^11^. Zhang et al.’s lung cancer risk model considered age, smoking status, lung functions, and hip/waist circumference. The CanPredict (lung) model developed by Liao et al. included sociodemographic, lifestyle, and medical history factors. Cassidy et al. ^12^ developed a model to estimate the 5-year probability of an individual developing lung cancer based on smoking duration, family history of lung cancer, prior diagnosis of pneumonia or other cancers, and occupational exposure to asbestos. Advancements in deep learning technologies have spurred interest in developing lung cancer risk models based on LDCT scans. Mikhael et al. ^13^ developed a model called Sybil to predict individual risk of developing lung cancer within six years based on LDCT, which relied on the presence of lung nodules and used bounding boxes to manually annotate the nodules. Notably, Sybil’s performance declined significantly when nodules were not included. Robbins et al. ^14^ proposed a risk-tailored approach for managing lung cancer screening results by incorporating individual risk factors and LDCT image features. Their model used both nodule features and non-nodule features (e.g., emphysema) to predict immediate and next-screen (1-year) lung cancer risks following both negative and abnormal LDCT results.

This study aimed to identify novel image biomarkers from LDCT chest scans that might signal the risk of developing lung cancer. Five different types of tissue related to body composition were segmented on LDCT images, which included subcutaneous adipose tissue (SAT), visceral adipose tissue (VAT), intramuscular adipose tissue (IMAT), skeletal muscle (SM), and bone. Volume and density metrics were computed for each tissue. Competing risk time-to-event analysis was used to identify body composition features significantly associated with the development of lung cancer and assess their hazard ratios. The underlying rationale for this approach is that body composition, which faithfully reflects an individual’s long-term habits and lifestyle (e.g., physical activity, exercise, and diet), provides the conditions that may promote or diminish cancer development. Body composition itself may provide a fertile global environment for cancer development. A detailed and in-depth study of various body tissues may gain unique insights into the role of body composition in lung cancer development and progression. The significant body composition features were combined with patient demographics (e.g., age, gender, race) to create a computer model to predict an individual’s risk of developing lung cancer, which was independent of the presence of suspicious nodules.

## MATERIALS AND METHODS

### The Pittsburgh Lung Screening Study (PLuSS)^15^

PLuSS is a community-based research cohort that screened and followed 3,635 current and ex-smokers using LDCT scans starting in 2002^15^ (Supplementary Table 1). Participants were enrolled between 2002 and 2005. The inclusion criteria were: (1) 50-79 years old and (2) current or ex-cigarette smokers with at least 12.5 pack-years at the time of enrollment. The exclusion criteria were: 1) quit smoking more than 10 years earlier, 2) had history of lung cancer diagnosis, or 3) had chest CT within one year prior to enrollment. Demographic, clinical, and smoking history data were collected using structured interviews and questionnaires at baseline and at annual follow-up visits or contacts. Upon study entry, participants underwent spirometry and LDCT screening and provided a sputum sample and a blood sample. A detailed description of the LDCT acquisition protocols can be found elsewhere ^15^. The cohort consisted of 1,869 men and 1,766 women, 94.1% of whom were white, 5.6% black, and 0.3% other non-white race. The mean age at enrollment was 59.1 years, and 60.2% were current smokers. Approximately 10% had a history of COPD and 20.6% had a family history of lung cancer involving a parent or sibling. This study has obtained ethics approval from the University of Pittsburgh Institutional Review Board (IRB 21020128).

As of February 2024, 1,463 (40.2%) participants in the PLuSS cohort died and 457 (12.6%) participants developed lung cancer, which included 340 patients who subsequently died. The follow-up time from the baseline ranges from 0.03 to 21.9 years with a median of 19.4 years. Among 1,463 participants who died, 9.4% died within 4 years, 25.4% within 8 years, and 47.2% within 12 years. Among 457 participants who had developed lung cancer, 104 (22.8%) developed cancer within the first 4 years, 172 (37.6%) developed cancer after 12 years.

### NLST-American College of Radiology Imaging Network (NLST-ACRIN) cohort

The NLST-ACRIN cohort includes 18,714 participants enrolled at 23 different centers of which 9,357 participants were randomized to annual LDCT screening^3, 16,17^. The inclusion criteria were: (1) Age 55-74 years; (2) 30 or more pack-year smoking history; and (3) quit smoking within the previous 15 years. The exclusion criteria were: (1) history of lung or other cancer; (2) history of lung surgery; and (3) chest CT examination in the 18 months prior to eligibility assessment. More detailed information about the clinical trial can be found at clinicaltrials.gov^17^. We have requested and received a sub-cohort (n=16,435) from NLST-ACRIN participants in the LDCT arm with demographic, clinical, and lung function data (Supplementary Table 2). The NLST CT scans were acquired using different scanners and protocols. The NLST-ACRIN participants were followed for 8 years (from 2002 to 2009) after the baseline visit. Notably, significant difference in the field-of-view (FOV) was observed for chest imaging between the NLST-ACRIN and PLuSS cohorts. Specifically, as shown in Supplementary Figure 1, approximately 15.8% of the baseline CT scans in the NLST-ACRIN cohort capture the entire chest FOV, while about 66.8% of the baseline CT scans in the PLuSS cohort provide a complete chest FOV. Additionally, roughly 90% of the baseline CT scans in the PLuSS cohort cover almost the entire chest, missing only limited regions. Based on these observations, we identified the CT scans in the NLST cohort with a complete chest FOV (n=2,604) as a separate test set (Supplementary Table 3) to evaluate the impact of chest FOV on the final performance. The criterion for determining whether the entire chest region is captured is to assess if the axial image slice containing the heart’s center includes the full chest region. The extent of the FOV is measured by counting the number of pixels of the chest region located at the image boundaries, with a value of zero indicating that the entire chest region is fully captured.

### LDCT image features

Artificial intelligence (AI) algorithms^18^ were used to automatically segment five types of tissues depicted on the LDCT in the PLuSS and NLST-ACRIN cohorts, including SAT, VAT, IMAT, SM, and bone. The volumes and densities (CT Hounsfield Unit) of the tissues were computed ^19^. Additionally, the ratios of each type of fat relative to the total body volume were computed.

### Statistical analysis and prediction modeling

All participants of both PLuSS and NLST cohort were included in the statistical analysis. The LDCT series with the maximum number of image slices from the baseline scans were used for analysis, prediction modeling, and validation.

The software “R” was used to analyze the risk from baseline LDCT scan to the diagnosis of lung cancer, accounting for death as a competing event. This competing risk time-to-event analysis was conducted to evaluate two groups of variables, including patient demographics and LDCT-derived body composition metrics, as predictors of future lung cancer. To ensure robustness, continuous parameters were standardized (mean zero, variance of one) for hazard ratio (HR) evaluation and effect comparison. A univariate analysis was performed first followed by joint modeling within each group. The final stage was a composite model integrating all variables from the two groups. HRs and their 95% confidence intervals (CIs) were computed to assess individual significance in univariate and combined models.

Two strategies were used to develop and validate the composite prediction models. The first strategy trained the risk prediction model using the PLuSS cohort and validated it using the NLST-ACRIN cohort or “PLuSS Model”. The second strategy trained the prediction model using 90% of both the PLuSS and NLST-ACRIN cohorts and validated it using the remaining 10% of each cohort or “MIX Model”. To ensure the robustness and reliability of the evaluation, the 10-fold cross-validation method was used. Considering that the maximum follow-up time in the NLST cohort is only 8 years, we cut off the follow-up time of the PLuSS cohort to 8 years to match this. Based on the above two training strategies, we trained another two models, namely “PLuSS_CUTOFF8 Model” and “MIX_CUTOFF8 Model.” The area under the curve (AUC) for standard time-dependent receiver operating characteristic (ROC) analysis measured the prognostic ability of the PLuSS and MIX models over the considered time range. The risk thresholds corresponding to specific sensitivity and specificity levels on a 12-year ROC curve were used to define the risk strata by grouping those with risks lower than the threshold at sensitivity = 0.9 as “low risk”, those with risks higher than the threshold at specificity = 0.9 as “high risk”, and the remaining as “intermediate risk”.

## RESULTS

### Demographics

Several demographic variables were significantly associated with lung cancer development in the univariate analysis (Table 1). Participants with active tobacco use (HR=1.73) and older participants (HR=1.57) had a significantly increased risk of developing lung cancer. Females (HR=0.75) and participants with a higher BMI (HR=0.84) had a significantly decreased risk of developing lung cancer. All univariately significant variables, except race, remained statistically significant in the demographic joint model. A model including gender, age, smoking status, and BMI achieved an AUC of 0.71 (95% CI: 0.67-0.74) for predicting the development of lung cancer within 12 years after the baseline CT scans (Fig. 1(a)). The performance of the demographic model to predict lung cancer development was reasonably consistent across the 21-year time period, except for the slight decrease in the beginning (Fig. 1(b)). The demographic joint model classified 933 (25.7%), 2,289 (63.0%), and 413 (11.4%) subjects into the low-risk, intermediate-risk, and high-risk strata, respectively (Table 2). Ninety-one high-risk subjects developed lung cancer within 12 years. The cumulative incidence of developing lung cancer for the three strata were clearly and evenly separated based on the demographic model (Fig. 2a).

**Figure 1.**
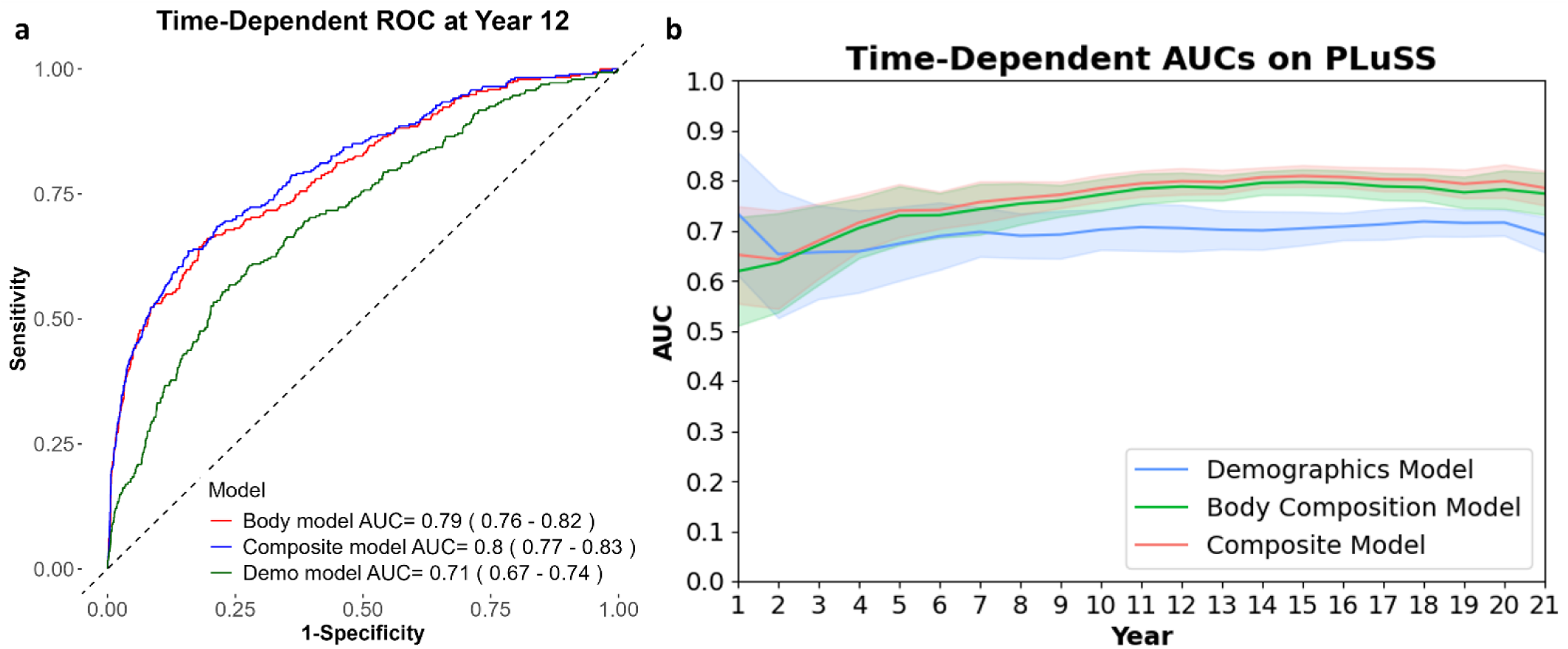
(a) ROC curves of demographics joint model, body composition joint model, and composite model predicting the development of lung cancer at 12 years in the PLuSS cohort. (b) Time-dependent AUCs of the joint models based on demographic variables, body composition variables, and composite model based on variables from two groups over 21 years tested on the PLuSS cohort.

**Figure 2.**
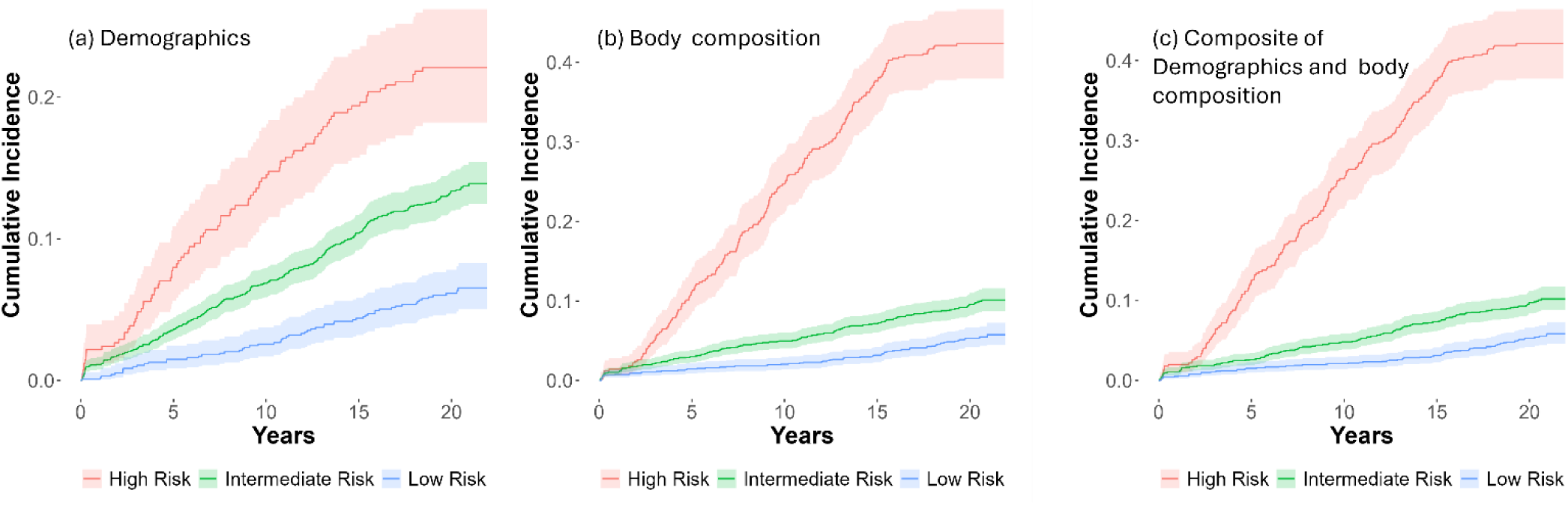
Cumulative incidences of lung cancer from baseline CT scans in the PLuSS cohort, categorized by low, intermediate, and high-risk groups. Risk stratification based on: (a) demographics, (b) body composition, and (c) the combination of demographics and body composition.

**Figure 3.**
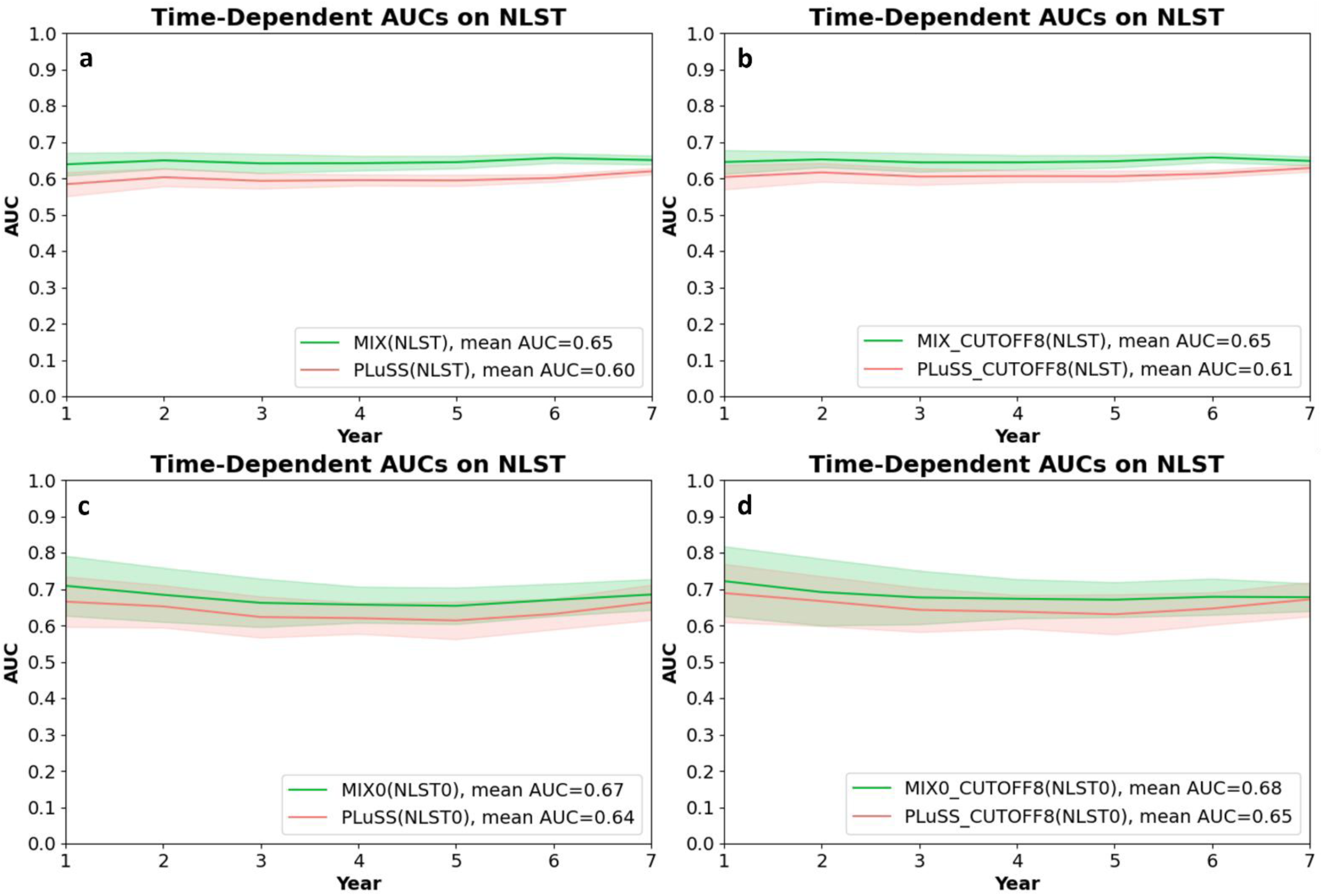
Time-dependent ROC curves of the PLuSS Model and the MIX Model. (a) PLuSS and MIX models tested on the NLST cohort. (b) PLuSS and MIX models with an 8-year follow-up cutoff tested on the NLST cohort. (c) PLuSS and MIX models trained on full FOV NLST scans and tested on full FOV NLST scans (10-fold cross-validation). (d) PLuSS and MIX models trained on full FOV NLST scans with an 8-year follow-up cutoff and tested on full FOV NLST scans. “MIX0” and “NLST0” refer to cases with a full chest FOV.

**Table 1.** Subject characteristics and body composition metrics’ association with lung cancer development (n=3,635)

| Variables | Univariate Cox Analysis |  | Joint Model |  |
| --- | --- | --- | --- | --- |
|  | Hazard Ratio | P Value | Hazard Ratio | P Value |
| **Demographics** |  |  |  |  |
| Gender (Female) | 0.75 | 0.0025 | 0.71 | 0.0004 |
| Age | 1.57 | <.0001 | 1.63 | <.0001 |
| Smoking status (Current) | 1.73 | <.0001 | 1.90 | <.0001 |
| Race (White) | 1.04 | 0.8548 | - | - |
| BMI | 0.84 | 0.0006 | 0.88 | 0.0160 |
| **Body composition** |  |  |  |  |
| VAT volume | 1.04 | 0.3756 | - | - |
| VAT ratio | 1.07 | 0.1453 | - | - |
| VAT density | 1.08 | 0.108 | - | - |
| SAT volume | 0.88 | 0.0096 | 0.51 | <0.0001 |
| SAT ratio | 0.86 | 0.0013 | - | - |
| SAT density | 1.12 | 0.0097 | - | - |
| IMAT volume | 0.89 | 0.0255 | - | - |
| IMAT ratio | 0.87 | 0.0047 | 0.31 | <0.0001 |
| IMAT density | 0.95 | 0.2715 | 0.56 | <0.0001 |
| SM volume | 1.07 | 0.1672 | - | - |
| SM ratio | 1.08 | 0.0806 | - | - |
| SM density | 0.71 | <.0001 | 0.26 | <0.0001 |
| Bone volume | 1.34 | <.0001 | 1.85 | <0.0001 |
| Bone ratio | 1.30 | <.0001 | - | - |
| Bone density | 0.68 | <.0001 | - | - |

Definition of abbreviations: BMI=body mass index; VAT=visceral adipose tissue; SAT=subcutaneous adipose tissue; IMAT=intramuscular adipose tissue; SM=skeletal muscle.

**Table 2.** Risk stratification of the baseline subjects in the PLuSS cohort (n=3,635)

| Variables | Risk stratum | Lung | Deaths | Total patients | 12-year risk |  |
| --- | --- | --- | --- | --- | --- | --- |
|  |  | Cancers |  |  | estimate | 95% CI |
| Demographics | Low risk | 59 | 139 | 933 | 3.3% | (2.3%, 4.6%) |
|  | Intermediate | 307 | 743 | 2289 | 8.0% | (7.0%, 9.2%) |
|  | High Risk | 91 | 241 | 413 | 16% | (13%, 20%) |
|  | Total | 457 | 1123 | 3635 |  |  |
| Body composition | Low risk | 69 | 313 | 1275 | 2.4% | (1.7%, 3.4%) |
|  | Intermediate | 182 | 642 | 1865 | 5.6 % | (4.6%, 6.7%) |
|  | High Risk | 206 | 168 | 495 | 30 % | (26%, 34%) |
|  | Total | 457 | 1123 | 3635 |  |  |
| Demographics and body composition | Low risk | 73 | 296 | 1322 | 2.4% | (1.7%, 3.4%) |
|  | Intermediate | 181 | 634 | 1815 | 5.8% | (4.8%, 6.9%) |
|  | High Risk | 203 | 193 | 498 | 29% | (25%, 33%) |
|  | Total | 457 | 1123 | 3635 |  |  |

### Body composition

Nine body composition variables were significantly associated with the development of lung cancer (Table 1). Bone volume (HR=1.34), bone ratio (HR=1.30), SAT density (HR=1.12) were significantly and directly related to the development of lung cancer. While muscle density (HR=0.71), bone density (HR=0.68), SAT volume (HR=0.88), SAT ratio (HR=0.86), IMAT volume (HR=0.89), and IMAT ratio (HR=0.87) were significantly and inversely related to the development of lung cancer. SM density (HR=0.26, p<0.0001), bone volume (HR=1.85, p<0.0001), IMAT ratio (HR=0.31, p<0.0001), IMAT density (HR=0.56, p<0.0001), and SAT volume (HR=0.51, p<0.0001) were significant variables in the joint model to predict lung cancer (Table 1). The joint model achieved a 12-year AUC of 0.79 (95% CI: 0.76-0.82), which is significantly higher than the joint model based on demographics variables (Fig. 1). IMAT density, which has non-significant effect univariately, became significantly predictive in the joint model. The body composition joint model classified 1,275 (35.1%), 1,865 (51.3%), and 495 (13.6%) subjects into the low-risk, intermediate-risk, and high-risk strata, respectively (Table 2). Among high-risk subjects, 206 developed lung cancer in 12 years. The cumulative incidence of developing lung cancer for the three strata is illustrated in Figure 2. The cumulative incidence of developing lung cancer for the high-risk stratum clearly separated from the low and intermediate-risk strata using the body composition model (Fig. 2b).

### Composite model

The final composite model included seven variables that were significantly associated with developing lung cancer: age (HR=1.20), current smoking status (HR=1.59), bone volume (HR=1.79), SM density (HR=0.29), IMAT ratio (HR=0.33), IMAT density (HR=0.56), and SAT volume (HR=0.56) (Table 3). The composite model classified 1,322 (36.4%), 1,815 (49.9%), and 498 (13.7%) subjects into the low-risk, intermediate-risk, and high-risk strata, respectively, for developing lung cancer (Table 2). Among high-risk subjects, 203 developed lung cancer in 12 years. The cumulative incidence of developing lung cancer for high-risk stratum clearly separated from the low and intermediate-risk strata using the composite model (Fig. 2c).

**Table 3.** Subject characteristics and body composition variables included in the final composite models for predicting lung cancer (n=3,635).

| **Variable** | **Hazard Ratio** | ***P* Value** |
| --- | --- | --- |
| Age | 1.20 | 0.0005 |
| Smoking status (Current) | 1.59 | <0.0001 |
| SAT volume | 0.56 | <0.0001 |
| IMAT ratio | 0.33 | <0.0001 |
| IMAT density | 0.56 | <0.0001 |
| SM density | 0.29 | <0.0001 |
| Bone volume | 1.79 | <0.0001 |

Definition of abbreviations: VAT=visceral adipose tissue; SAT=subcutaneous adipose tissue; IMAT=intramuscular adipose tissue; SM=skeletal muscle.

The PLuSS Model based on the seven variables achieved a 12-year AUC of 0.80 (95% CI: 0.77-0.83), which showed only a marginal improvement compared to the body composition model (AUC of 0.79, 95% CI: 0.76-0.82). The likelihood ratio test indicated a statistically significant difference between the two models. Both the PLuSS Model and the body composition model were statistically better than the demographic model (AUC of 0.71, 95% CI: 0.67-0.74). The PLuSS Model achieved a mean AUC of 0.70 (95% CI: 0.66-0.74) on PLuSS data with 10-fold cross validation over the first 7 years. The mean AUC was 0.60 (95% CI: 0.59 - 0.61) over 7 years when validated on the baseline NLST LDCT scans, 0.64 (95% CI: 0.62-0.66) when validated on NLST scans with full chest FOV. The PLuSS_CUTOFF8 model achieved a mean AUC of 0.61 (95% CI: 0.60-0.62) when validated on all baseline NLST scans, and a mean AUC of 0.65 (95% CI: 0.64-0.67) when validated on baseline NLST scans with a full chest FOV.

In contrast, both the MIX Model and MIX_CUTOFF8 Model achieved a mean AUC of 0.65 (95% CI: 0.64-0.65) on all the cases in the NLST cohort over 7 years based on 10-fold cross-validation. The MIX Model achieved a mean AUC of 0.67 (95% CI: 0.66-0.69) on NLST scans with full chest FOV. The MIX_CUTOFF8 Model achieved a mean AUC of 0.68 (95% CI: 0.67-0.70) on NLST scans with full chest FOV.

## DISCUSSION

The comprehensive analysis of the body composition revealed that both body composition and subject characteristics are associated with the risk of developing lung cancer. Our promising results from two LDCT screening cohorts demonstrate that body composition measures are significantly associated with lung cancer risk and can be used to stratify subjects into three risk strata. All types of tissues related to body composition analyzed (i.e., adipose, bone, and muscle) were significant predictors of lung cancer development and able to stratify subjects into three risk categories. Despite the significance of the demographic information in the model, their inclusion does not improve the performance of the model based on body composition very much in terms of AUC (Fig. 1 and Table 2), suggesting that the characteristics of body composition may imply patient information (e.g., age and gender). The ability to assess an individual’s risk of developing lung cancer can lead to a personalized lung cancer screening strategy. Individuals with a lower risk of developing lung cancer could undergo less frequent LDCT-based screening (e.g., every 2-3 years), while those at high risk would maintain annual screenings. A personalized screening strategy may substantially reduce unnecessary radiation exposure associated with LDCT imaging and other potentially harmful procedures (e.g., lung biopsies). Ultimately, these factors could help reduce the healthcare burden of lung cancer and enable the implementation of proactive preventative measures.

The performance of the models for predicting lung cancer in the PLuSS cohort was promising and showed an upward trend over time (Fig. 1). In the first five years, the models’ performance was relatively lower, with an AUC below 0.70. However, the performance improved, reaching an AUC of 0.81 around 15 years, before starting to slightly decline around 16 years. When the PLuSS model was directly applied to cases in the NLST-ACRIN cohort, the prediction performance, measured by AUC over the 7 follow-up years, was approximately 0.60-0.61. This was consistent whether or not the follow-up period in the PLuSS cohort was truncated to match that of the NLST-ACRIN cohort. Similarly, when the MIX model was applied to the NLST-ACRIN cohort, the AUC remained around 0.65, regardless of the follow-up period adjustment. Furthermore, when testing only cases with a full chest FOV in the NLST-ACRIN cohort, the prediction performance improved to 0.64-0.65 for the PLuSS model and 0.67-0.68 for the MIX model. In fact, this performance is similar to that of the PLuSS model for predicting lung cancer over the first 7 years (Fig. 1). These results suggest that the chest FOV and potential imaging protocols significantly impact prediction accuracy. A complete FOV is associated with improved performance. Additionally, using a diverse dataset, as in the MIX model, is also linked to enhanced prediction accuracy. Nevertheless, considering that the PLuSS cohort was followed for around 22 years while the NLST-ACRIN cohort was followed for only 8 years, along with their differences in image acquisition protocols, these results strongly support the potential association between body composition and lung cancer risk.

There has been significant interest in understanding the relationship between body composition and cancer, and a number of studies have been conducted to shed light on this topic^20, 21^. However, most of these studies have primarily focused on the use of BMI or waist circumference as measures of body composition. While some studies have found an inverse association between BMI and the development of lung cancer, there are also conflicting results. Some studies ^22–27^ have found an inverse association between BMI and lung cancer risk and some have found a positive association between waist circumference and lung cancer risk. Other studies^28, 29^ have failed to confirm this association and have even reported contradictory results^30^.BMI has been reported to be unrelated to lung cancer risk when limited to non-smokers^31^ ^32^. Therefore, it is crucial to clarify the association between BMI and the risk of lung cancer, as it has important implications for public health. One potential reason for the conflicting results in the literature may be that the simple BMI assessment does not adequately capture an individual’s body composition. BMI does not assess visceral fat, intramuscular fat, and bone density. These factors may differentially impact the development of lung cancer.

Xu et al. ^33^ conducted a secondary body composition analysis of the NLST that included area and attenuation attributes of SM and SAT on LDCT scans. They observed that body composition measurements added predictive value for lung cancer death, cardiovascular disease death, and all-cause death, but not for lung cancer incidence in NLST. Their conclusion about lung cancer differs from ours. This discrepancy could be due to differences in the body composition assessment. They only segmented SM and SAT on three CT image slices. Consequently, they quantified the area of these tissues instead of their volume, which reduced the “resolution” of the tissue assessment based on a limited number of images. To our knowledge, most of studies primarily focused on BMI ^22–27^ or only evaluated a few types of tissues based on a limited number of images at specific anatomical location (e.g., T4 or L3 vertebra) ^34–41^. In contrast, our study utilized AI algorithms to identify five different body tissues and quantify their volumetric measures using all the images in a CT scan.

At this time, we have not utilized chest CT scans directly as inputs to train a deep learning model for predictive purposes, despite extensive experience in this field ^42–47^. First, deep learning of this nature typically demands an extremely large dataset. While our cohorts are relatively large, they are insufficient to reliably train such a deep learning model due to the limited number of cancer cases and the inherent anatomical variability between individuals. Second, a deep learning model operates as a “black box,” which complicates the interpretation of factors associated with lung cancer risk. In contrast, an AI model based on manually crafted CT-derived features offers greater interpretability regarding their connection to lung cancer risk, which is crucial for developing effective prevention strategies.

## CONCLUSION

Lung cancer screening using LDCT has proven effective in reducing mortality from lung cancer. However, there are challenges to screening such as low incidence among screened individuals and false positive detections and the risks associated with radiation exposure and follow-up procedures. Current screening criteria based solely on age and smoking history can tend to overestimate an individual’s lung cancer risk, which can be an issue in any type of mass screening. There incidence of cancer is low in the general population, which results in mostly negative screening results. This study unveils the significant associations between body tissues and lung cancer risk, which can help classify individuals into different risk strata. Implementing risk-stratified screening strategies based on these models could improve screening efficacy by identifying low-risk individuals who may benefit from reduced screening intervals based on more personalized lung cancer screening. A more personalized screening approach may help reduce the frequency of screening exams and the number of potentially harmful follow-up procedures (e.g., radiation exposure from additional CT scans, invasive biopsies) associated with false positive screening results.

## Abbreviations

AI: artificial intelligence

ACRIN: American College of Radiology Imaging Network

AUC: area under curve

BMI: body mass index

CI: confidence interval

FOV: field-of-view

HR: hazard ratio

IMAT: intramuscular adipose tissue

LDCT: low-dose computed tomography

PLuSS: Pittsburgh Lung Screening Study

NLST: National Lung Screening Trial

ROC: receiver operating characteristic

SAT: subcutaneous adipose tissue

SM: skeletal muscle

VAT: visceral adipose tissue

## Data Availability

All data produced in the present study are available upon reasonable request to the authors

## Acknowledgements

This work is supported in part by research grants from the National Institutes of Health (NIH) (R01CA237277, U01CA271888, and P30CA047904) and UPMC Hillman Developmental Pilot Program.

## REFERENCES

1. Cancer Facts & Figures 2019 2019. Available from: https://www.cancer.org/research/cancer-facts-statistics/all-cancer-facts-figures/cancer-facts-figures-2019.html.

2. Aberle DR, DeMello S, Berg CD, Black WC, Brewer B, Church TR, Clingan KL, Duan F, Fagerstrom RM, Gareen IF, Gatsonis CA, Gierada DS, Jain A, Jones GC, Mahon I, Marcus PM, Rathmell JM, Sicks J, National Lung Screening Trial Research T. Results of the two incidence screenings in the National Lung Screening Trial. N Engl J Med. 2013;369(10):920–31. Epub 2013/09/06. doi: 10.1056/NEJMoa1208962. PubMed PMID: 24004119; PMCID: PMC4307922.

3. National Lung Screening Trial Research T, Aberle DR, Adams AM, Berg CD, Black WC, Clapp JD, Fagerstrom RM, Gareen IF, Gatsonis C, Marcus PM, Sicks JD. Reduced lung-cancer mortality with low-dose computed tomographic screening. N Engl J Med. 2011;365(5):395–409. Epub 2011/07/01. doi: 10.1056/NEJMoa1102873. PubMed PMID: 21714641; PMCID: PMC4356534.

4. National Lung Screening Trial Research T, Aberle DR, Berg CD, Black WC, Church TR, Fagerstrom RM, Galen B, Gareen IF, Gatsonis C, Goldin J, Gohagan JK, Hillman B, Jaffe C, Kramer BS, Lynch D, Marcus PM, Schnall M, Sullivan DC, Sullivan D, Zylak CJ. The National Lung Screening Trial: overview and study design. Radiology. 2011;258(1):243–53. Epub 2010/11/04. doi: 10.1148/radiol.10091808. PubMed PMID: 21045183; PMCID: PMC3009383.

5. de Koning HJ, van der Aalst CM, de Jong PA, Scholten ET, Nackaerts K, Heuvelmans MA, Lammers JJ, Weenink C, Yousaf-Khan U, Horeweg N, van’t Westeinde S, Prokop M, Mali WP, Mohamed Hoesein FAA, van Ooijen PMA, Aerts J, den Bakker MA, Thunnissen E, Verschakelen J, Vliegenthart R, Walter JE, Ten Haaf K, Groen HJM, Oudkerk M. Reduced Lung-Cancer Mortality with Volume CT Screening in a Randomized Trial. N Engl J Med. 2020;382(6):503–13. Epub 2020/01/30. doi: 10.1056/NEJMoa1911793. PubMed PMID: 31995683.

6. Jin J. Screening for Lung Cancer. JAMA. 2021;325(10):1016. Epub 2021/03/10. doi: 10.1001/jama.2021.1799. PubMed PMID: 33687464.

7. Ardila D, Kiraly AP, Bharadwaj S, Choi B, Reicher JJ, Peng L, Tse D, Etemadi M, Ye W, Corrado G, Naidich DP, Shetty S. End-to-end lung cancer screening with three-dimensional deep learning on low-dose chest computed tomography. Nat Med. 2019;25(6):954–61. Epub 2019/05/22. doi: 10.1038/s41591-019-0447-x. PubMed PMID: 31110349.

8. Ashraf SF, Yin K, Meng CX, Wang Q, Wang Q, Pu J, Dhupar R. Predicting benign, preinvasive, and invasive lung nodules on computed tomography scans using machine learning. J Thorac Cardiovasc Surg. 2022;163(4):1496–505 e10. Epub 2021/03/18. doi: 10.1016/j.jtcvs.2021.02.010. PubMed PMID: 33726909.

9. Raghu VK, Zhao W, Pu J, Leader JK, Wang R, Herman J, Yuan JM, Benos PV, Wilson DO. Feasibility of lung cancer prediction from low-dose CT scan and smoking factors using causal models. Thorax. 2019;74(7):643–9. Epub 2019/03/14. doi: 10.1136/thoraxjnl-2018-212638. PubMed PMID: 30862725; PMCID: PMC6585306.

10. Zhang S, Yang L, Xu W, Wang Y, Han L, Zhao G, Cai T. Predicting the risk of lung cancer using machine learning: A large study based on UK Biobank. Medicine (Baltimore). 2024;103(16):e37879. Epub 2024/04/19. doi: 10.1097/MD.0000000000037879. PubMed PMID: 38640268; PMCID: PMC11029993.

11. Liao W, Coupland CAC, Burchardt J, Baldwin DR, initiative D, Gleeson FV, Hippisley-Cox J. Predicting the future risk of lung cancer: development, and internal and external validation of the CanPredict (lung) model in 19.67 million people and evaluation of model performance against seven other risk prediction models. Lancet Respir Med. 2023;11(8):685–97. Epub 2023/04/09. doi: 10.1016/S2213-2600(23)00050-4. PubMed PMID: 37030308.

12. Cassidy A, Myles JP, van Tongeren M, Page RD, Liloglou T, Duffy SW, Field JK. The LLP risk model: an individual risk prediction model for lung cancer. Br J Cancer. 2008;98(2):270–6. Epub 2007/12/19. doi: 10.1038/sj.bjc.6604158. PubMed PMID: 18087271; PMCID: PMC2361453.

13. Mikhael PG, Wohlwend J, Yala A, Karstens L, Xiang J, Takigami AK, Bourgouin PP, Chan P, Mrah S, Amayri W, Juan YH, Yang CT, Wan YL, Lin G, Sequist LV, Fintelmann FJ, Barzilay R. Sybil: A Validated Deep Learning Model to Predict Future Lung Cancer Risk From a Single Low-Dose Chest Computed Tomography. J Clin Oncol. 2023;41(12):2191–200. Epub 2023/01/13. doi: 10.1200/JCO.22.01345. PubMed PMID: 36634294; PMCID: PMC10419602 manuscript.

14. Robbins HA, Cheung LC, Chaturvedi AK, Baldwin DR, Berg CD, Katki HA. Management of Lung Cancer Screening Results Based on Individual Prediction of Current and Future Lung Cancer Risks. J Thorac Oncol. 2022;17(2):252–63. Epub 2021/10/15. doi: 10.1016/j.jtho.2021.10.001. PubMed PMID: 34648946; PMCID: PMC10186153.

15. Wilson DO, Weissfeld JL, Fuhrman CR, Fisher SN, Balogh P, Landreneau RJ, Luketich JD, Siegfried JM. The Pittsburgh Lung Screening Study (PLuSS): outcomes within 3 years of a first computed tomography scan. Am J Respir Crit Care Med. 2008;178(9):956–61. Epub 2008/07/19. doi: 10.1164/rccm.200802-336OC. PubMed PMID: 18635890; PMCID: PMC2720144.

16. Young RP, Duan F, Chiles C, Hopkins RJ, Gamble GD, Greco EM, Gatsonis C, Aberle D. Airflow Limitation and Histology Shift in the National Lung Screening Trial. The NLST-ACRIN Cohort Substudy. American journal of respiratory and critical care medicine. 2015;192(9):1060–7. doi: 10.1164/rccm.201505-0894OC. PubMed PMID: 26199983; PMCID: PMC4642202.

17. National Lung Screening Trial (NLST) Screening (NLST) [cited 2023 September 1]. Available from: https://classic.clinicaltrials.gov/ct2/show/NCT00047385.

18. Pu L, Gezer NS, Ashraf SF, Ocak I, Dresser DE, Dhupar R. Automated segmentation of five different body tissues on computed tomography using deep learning. Med Phys. 2023;50(1):178–91. Epub 2022/08/26. doi: 10.1002/mp.15932. PubMed PMID: 36008356.

19. Pu L, Ashraf SF, Gezer NS, Ocak I, Dresser DE, Leader JK, Dhupar R. Estimating 3-D whole-body composition from a chest CT scan. Med Phys. 2022;49(11):7108–17. Epub 2022/06/24. doi: 10.1002/mp.15821. PubMed PMID: 35737963.

20. He Q, Xia B, Liu A, Li M, Zhou Z, Cheung EC, Kuo ZC, Wang B, Li F, Tang Y, Zheng Z, Sun R, Hu YJ, Meng W, He Y, Yuan J, Zhang C. Association of body composition with risk of overall and site-specific cancers: A population-based prospective cohort study. Int J Cancer. 2021;149(7):1435–47. Epub 2021/05/22. doi: 10.1002/ijc.33697. PubMed PMID: 34019699.

21. Jeong SM, Lee DH, Giovannucci EL. Predicted lean body mass, fat mass and risk of lung cancer: prospective US cohort study. Eur J Epidemiol. 2019;34(12):1151–60. Epub 2019/11/23. doi: 10.1007/s10654-019-00587-2. PubMed PMID: 31754943; PMCID: PMC7504685.

22. Dewi NU, Boshuizen HC, Johansson M, Vineis P, Kampman E, Steffen A, Tjonneland A, Halkjaer J, Overvad K, Severi G, Fagherazzi G, Boutron-Ruault MC, Kaaks R, Li K, Boeing H, Trichopoulou A, Bamia C, Klinaki E, Tumino R, Palli D, Mattiello A, Tagliabue G, Peeters PH, Vermeulen R, Weiderpass E, Torhild Gram I, Huerta JM, Agudo A, Sanchez MJ, Ardanaz E, Dorronsoro M, Quiros JR, Sonestedt E, Johansson M, Grankvist K, Key T, Khaw KT, Wareham N, Cross AJ, Norat T, Riboli E, Fanidi A, Muller D, Bueno-de-Mesquita HB. Anthropometry and the Risk of Lung Cancer in EPIC. Am J Epidemiol. 2016;184(2):129–39. Epub 2016/07/03. doi: 10.1093/aje/kwv298. PubMed PMID: 27370791; PMCID: PMC4945700.

23. Yu D, Zheng W, Johansson M, Lan Q, Park Y, White E, Matthews CE, Sawada N, Gao YT, Robien K, Sinha R, Langhammer A, Kaaks R, Giovannucci EL, Liao LM, Xiang YB, Lazovich D, Peters U, Zhang X, Bueno-de-Mesquita B, Willett WC, Tsugane S, Takata Y, Smith-Warner SA, Blot W, Shu XO. Overall and Central Obesity and Risk of Lung Cancer: A Pooled Analysis. J Natl Cancer Inst. 2018;110(8):831–42. Epub 2018/03/09. doi: 10.1093/jnci/djx286. PubMed PMID: 29518203; PMCID: PMC6093439.

24. Ardesch FH, Ruiter R, Mulder M, Lahousse L, Stricker BHC, Kiefte-de Jong JC. The Obesity Paradox in Lung Cancer: Associations With Body Size Versus Body Shape. Front Oncol. 2020;10:591110. Epub 2020/11/28. doi: 10.3389/fonc.2020.591110. PubMed PMID: 33244459; PMCID: PMC7683800.

25. Kabat GC, Kim M, Hunt JR, Chlebowski RT, Rohan TE. Body mass index and waist circumference in relation to lung cancer risk in the Women’s Health Initiative. Am J Epidemiol. 2008;168(2):158–69. Epub 2008/05/17. doi: 10.1093/aje/kwn109. PubMed PMID: 18483121; PMCID: PMC2878097.

26. Smith L, Brinton LA, Spitz MR, Lam TK, Park Y, Hollenbeck AR, Freedman ND, Gierach GL. Body mass index and risk of lung cancer among never, former, and current smokers. J Natl Cancer Inst. 2012;104(10):778–89. Epub 2012/03/30. doi: 10.1093/jnci/djs179. PubMed PMID: 22457475; PMCID: PMC3352831.

27. Tarnaud C, Guida F, Papadopoulos A, Cenee S, Cyr D, Schmaus A, Radoi L, Paget-Bailly S, Menvielle G, Buemi A, Woronoff AS, Luce D, Stucker I. Body mass index and lung cancer risk: results from the ICARE study, a large, population-based case-control study. Cancer Causes Control. 2012;23(7):1113–26. Epub 2012/05/23. doi: 10.1007/s10552-012-9980-3. PubMed PMID: 22610667.

28. Jee SH, Yun JE, Park EJ, Cho ER, Park IS, Sull JW, Ohrr H, Samet JM. Body mass index and cancer risk in Korean men and women. Int J Cancer. 2008;123(8):1892–6. Epub 2008/07/25. doi: 10.1002/ijc.23719. PubMed PMID: 18651571.

29. Parr CL, Batty GD, Lam TH, Barzi F, Fang X, Ho SC, Jee SH, Ansary-Moghaddam A, Jamrozik K, Ueshima H, Woodward M, Huxley RR, Asia-Pacific Cohort Studies C. Body-mass index and cancer mortality in the Asia-Pacific Cohort Studies Collaboration: pooled analyses of 424,519 participants. Lancet Oncol. 2010;11(8):741–52. Epub 2010/07/03. doi: 10.1016/S1470-2045(10)70141-8. PubMed PMID: 20594911; PMCID: PMC4170782.

30. Kabat GC, Miller AB, Rohan TE. Body mass index and lung cancer risk in women. Epidemiology. 2007;18(5):607–12. Epub 2007/09/20. doi: 10.1097/ede.0b013e31812713d1. PubMed PMID: 17879428.

31. Henley SJ, Flanders WD, Manatunga A, Thun MJ. Leanness and lung cancer risk: fact or artifact? Epidemiology. 2002;13(3):268–76. Epub 2002/04/20. doi: 10.1097/00001648-200205000-00006. PubMed PMID: 11964927.

32. Lam TK, Moore SC, Brinton LA, Smith L, Hollenbeck AR, Gierach GL, Freedman ND. Anthropometric measures and physical activity and the risk of lung cancer in never-smokers: a prospective cohort study. PLoS One. 2013;8(8):e70672. Epub 2013/08/14. doi: 10.1371/journal.pone.0070672. PubMed PMID: 23940620; PMCID: PMC3734257.

33. Xu K, Khan MS, Li TZ, Gao R, Terry JG, Huo Y, Lasko TA, Carr JJ, Maldonado F, Landman BA, Sandler KL. AI Body Composition in Lung Cancer Screening: Added Value Beyond Lung Cancer Detection. Radiology. 2023;308(1):e222937. Epub 2023/07/25. doi: 10.1148/radiol.222937. PubMed PMID: 37489991; PMCID: PMC10374937.

34. Dabiri S, Popuri K, Cespedes Feliciano EM, Caan BJ, Baracos VE, Beg MF. Muscle segmentation in axial computed tomography (CT) images at the lumbar (L3) and thoracic (T4) levels for body composition analysis. Comput Med Imaging Graph. 2019;75:47–55. Epub 2019/05/28. doi: 10.1016/j.compmedimag.2019.04.007. PubMed PMID: 31132616; PMCID: PMC6620151.

35. Nowak S, Faron A, Luetkens JA, Geissler HL, Praktiknjo M, Block W, Thomas D, Sprinkart AM. Fully Automated Segmentation of Connective Tissue Compartments for CT-Based Body Composition Analysis: A Deep Learning Approach. Invest Radiol. 2020;55(6):357–66. Epub 2020/05/06. doi: 10.1097/RLI.0000000000000647. PubMed PMID: 32369318.

36. Lee H, Troschel FM, Tajmir S, Fuchs G, Mario J, Fintelmann FJ, Do S. Pixel-Level Deep Segmentation: Artificial Intelligence Quantifies Muscle on Computed Tomography for Body Morphometric Analysis. J Digit Imaging. 2017;30(4):487–98. Epub 2017/06/28. doi: 10.1007/s10278-017-9988-z. PubMed PMID: 28653123; PMCID: PMC5537099.

37. Wang Y, Qiu Y, Thai T, Moore K, Liu H, Zheng B. A two-step convolutional neural network based computer-aided detection scheme for automatically segmenting adipose tissue volume depicting on CT images. Comput Methods Programs Biomed. 2017;144:97–104. Epub 2017/05/13. doi: 10.1016/j.cmpb.2017.03.017. PubMed PMID: 28495009; PMCID: PMC5441239.

38. Bridge CP, Rosenthal M, Wright B, Kotecha G, Fintelmann F, Troschel F, Miskin N, Desai K, Wrobel W, Babic A, Khalaf N, Brais L, Welch M, Zellers C, Tenenholtz N, Michalski M, Wolpin B, Andriole K, editors. Fully-Automated Analysis of Body Composition from CT in Cancer Patients Using Convolutional Neural Networks. OR

20 Context-Aware Operating Theaters, Computer Assisted Robotic Endoscopy, Clinical Image-Based Procedures, and Skin Image Analysis; 2018 2018//; Cham: Springer International Publishing.

39. Weston AD, Korfiatis P, Kline TL, Philbrick KA, Kostandy P, Sakinis T, Sugimoto M, Takahashi N, Erickson BJ. Automated Abdominal Segmentation of CT Scans for Body Composition Analysis Using Deep Learning. Radiology. 2019;290(3):669–79. Epub 2018/12/12. doi: 10.1148/radiol.2018181432. PubMed PMID: 30526356.

40. Koitka S, Kroll L, Malamutmann E, Oezcelik A, Nensa F. Fully automated body composition analysis in routine CT imaging using 3D semantic segmentation convolutional neural networks. Eur Radiol. 2021;31(4):1795–804. Epub 2020/09/19. doi: 10.1007/s00330-020-07147-3. PubMed PMID: 32945971; PMCID: PMC7979624.

41. Huber FA, Chaitanya K, Gross N, Chinnareddy SR, Gross F, Konukoglu E, Guggenberger R. Whole-body Composition Profiling Using a Deep Learning Algorithm: Influence of Different Acquisition Parameters on Algorithm Performance and Robustness. Invest Radiol. 2021. Epub 2021/06/03. doi: 10.1097/RLI.0000000000000799. PubMed PMID: 34074943.

42. Beeche C, Singh JP, Leader JK, Gezer NS, Oruwari1 AP, Dansingani KK, Chhablani J, Pu J. Super U-Net: a modularized generalizable architecture. Pattern Recognition. 2022;128(2):108669.

43. Pu J, Leader JK, Sechrist J, Beeche CA, Singh JP, Ocak IK, Risbano MG. Automated identification of pulmonary arteries and veins depicted in non-contrast chest CT scans. Med Image Anal. 2022;77:102367. Epub 2022/01/24. doi: 10.1016/j.media.2022.102367. PubMed PMID: 35066393; PMCID: PMC8901546.

44. Wang L, Chen K, Wen H, Zheng Q, Chen Y, Pu J, Chen W. Feasibility assessment of infectious keratitis depicted on slit-lamp and smartphone photographs using deep learning. International Journal of Medical Informatics. 2021;155:104583. doi: 10.1016/j.ijmedinf.2021.104583.

45. Wang L, Gu J, Chen Y, Liang Y, Zhang W, Pu J, Chen H. Automated segmentation of the optic disc from fundus images using an asymmetric deep learning network. Pattern Recognit. 2021;112. Epub 2021/08/07. doi: 10.1016/j.patcog.2020.107810. PubMed PMID: 34354302; PMCID: PMC8336919.

46. Ashraf SF, Yin K, Meng CX, Wang Q, Wang Q, Pu J, Dhupar R. Predicting benign, preinvasive, and invasive lung nodules on computed tomography scans using machine learning. J Thorac Cardiovasc Surg. 2021. Epub 2021/03/18. doi: 10.1016/j.jtcvs.2021.02.010. PubMed PMID: 33726909.

47. Wang X, Yu J, Zhu Q, Li S, Zhao Z, Yang B, Pu J. Potential of deep learning in assessing pneumoconiosis depicted on digital chest radiography. Occupational and Environmental Medicine. 2020:oemed-2019-106386. doi: 10.1136/oemed-2019-106386.

