## Supplemental Table 1-3, Figure 1 for "Body composition as a biomarker for assessing future lung cancer risk"

**Supplementary Material**

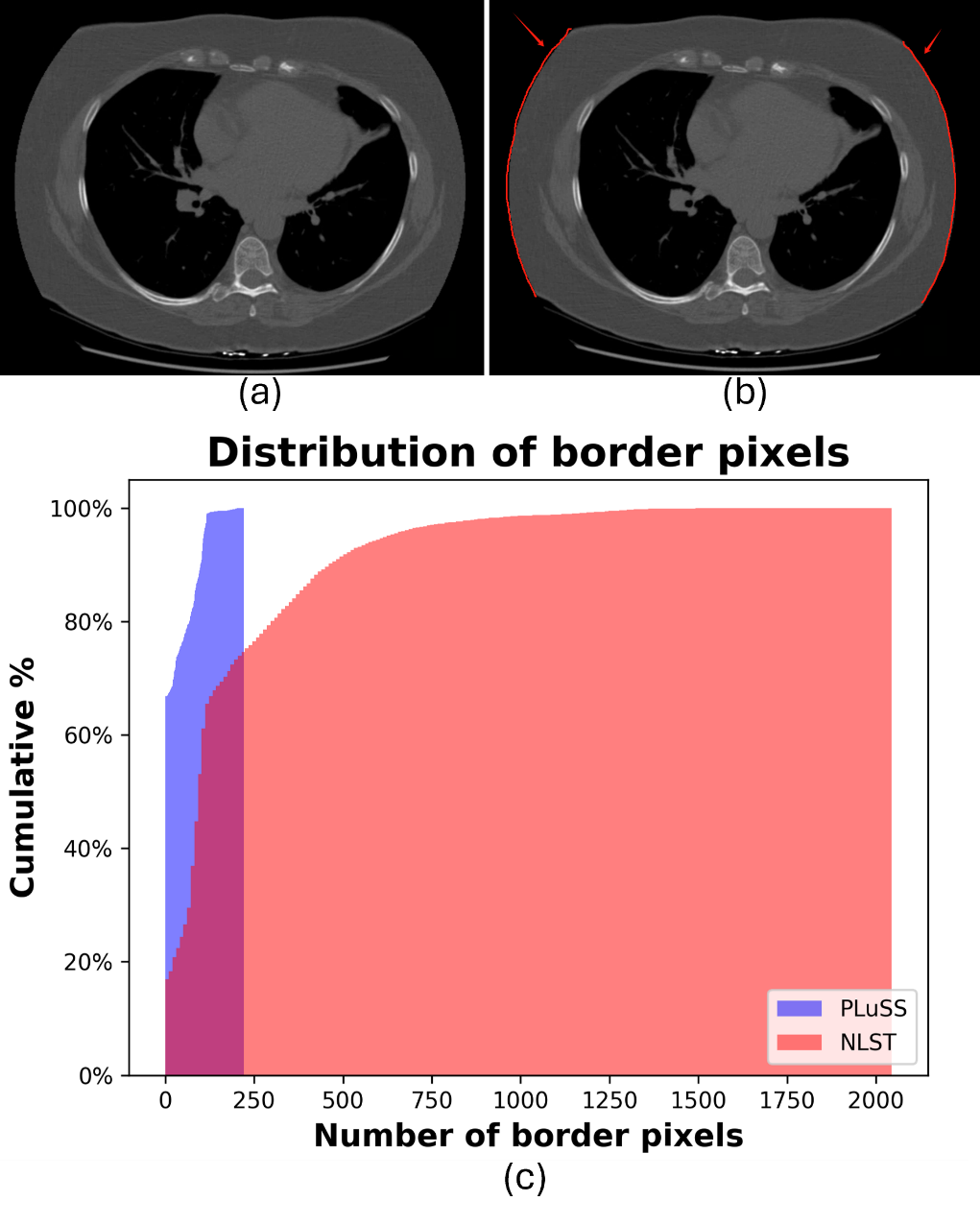

**Supplementary Figure 1**. Completeness of the Chest Field-of-View (FOV) in the PLuSS and NLST-ACRIN Cohorts. (a) A CT image slice showing the location of the heart center. (b) The chest region at the image boundaries, indicated by red lines. (c) The percentage of cases with incomplete chest regions, based on the number of border pixels

**Supplementary Table 1:** Summary of the Pittsburgh Lung Screening Study (PLuSS) cohort (n=3,635)

| **Demographics** | **Lung Cancer Incidence (*n*=457)** | **Death Incidence (*n*=1,463)** | **Event-free Incidence (*n*=2,055)** | **All Participants (*n*=3,635)** |
| --- | --- | --- | --- | --- |
| Gender (Female) | 199 (43.5) * | 597 (40.8) * | 1110 (54.0) | 1,766 (48.6) * |
| Age (years) | 61.0 (55.0-66.0) * | 63.0 (57.0-68.0) * | 56.0 (53.0-60.0) | 58.0 (54.0-64.0) * |
| Smoker | 326 (71.3) * | 926 (63.3) * | 1180 (57.4) | 2,188 (60.2) * |
| Race (White) | 432 (94.5) | 1380 (94.3) | 1928 (93.8) | 3,422 (94.1) |
| BMI | 27.4 (24.5-31.3) * | 28.1 (25.0-32.1) | 28.0 (25.2-31.6) | 28.0 (25.1-31.8) |
| Definition of abbreviations: BMI=body mass index  *p < 0.05 compared with event-free group. | | | | |

**Supplementary Table 2:** Summary of the National Lung Screening Trial (NLST) cohort (n=16,435)

| **Demographics** | **Lung Cancer Incidence (*n*=1,028)** | **Death Incidence (*n*=1,212)** | **Event-free Incidence (*n*=14,664)** | **All Participants (*n*=16,435)** |
| --- | --- | --- | --- | --- |
| Gender (Female) | 408 (39.7) | 367 (30.3) * | 6091 (41.5) | 6708 (40.8) |
| Age (years) | 63.0 (59.0-68.0) * | 64.0 (59.0-68.0) * | 61.0 (57.0-65.0) | 61.0 (57.0-65.0) * |
| Smoker | 597 (58.1) * | 720 (59.4) * | 6853 (46.7) | 7879 (47.9) * |
| Race (White) | 944 (91.8) | 1101 (90.8) | 13496 (92.0) | 15106 (91.9) |
| BMI | 26.3 (23.7-29.2) * | 26.9 (23.7-30.6) * | 27.3 (24.4-30.5) | 27.2 (24.4-30.5) |
| Definition of abbreviations: BMI=body mass index  *p < 0.05 compared with event-free group. | | | | |

**Supplementary Table 3:** Summary of the cases with a complete chest field-of-view (FOV) in the National Lung Screening Trial (NLST) cohort (n=2,604)

| **Demographics** | **Lung Cancer Incidence (*n*=197)** | **Death Incidence (*n*=226)** | **Event-free Incidence (*n*=2,268)** | **All Participants (*n*=2,604)** |
| --- | --- | --- | --- | --- |
| Gender (Female), n (%) | 66 (33.5) | 57 (25.2) * | 885 (39.0) | 988 (37.9) |
| Age (years), mean (SD) | 64.5 (5.5) * | 64.7 (5.7) * | 61.9 (5.1) | 62.2 (5.3) * |
| Smoking status (Current), n (%) | 121 (61.4) | 149 (65.9) * | 1245 (54.9) | 1458 (56.0) |
| Race (White), n (%) | 186 (94.4) | 210 (92.9) | 2160 (95.2) | 2473 (95.0) |
| BMI, mean (SD) | 24.9 (3.8) * | 24.9 (4.3) * | 25.8 (4.2) | 25.7 (4.2) |
| Definition of abbreviations: BMI=body mass index  *p < 0.05 compared with event-free group. | | | | |
